# Trends and factors associated with trichomoniasis among people attending sexual health services, England, 2012 to 2023

**DOI:** 10.1101/2025.08.12.25333487

**Authors:** Ana Karina Harb, George Baldry, Stephanie J Migchelsen, Laura Viviani, Michelle Cole, Rachel Pitt-Kendall, Hamish Mohammed

## Abstract

**Objective:** We assessed trends and risk factors for trichomoniasis since the publication of the 2014 BASHH trichomoniasis management guideline and following the addition, in 2021, of recommended [demographically and geographically] targeted screening for this infection.

**Methods:** We described gender-stratified trends in trichomoniasis diagnoses at sexual health services in England from 2012-2023 using data from the GUMCAD STI surveillance system. We determined gender-stratified factors associated with trichomoniasis diagnosis.

**Results:** Nationally, trichomoniasis diagnoses increased by 38% between 2012 and 2019 (6,950 to 9,490 diagnoses); after a drop in 2020 due to COVID-19, diagnoses increased by 54% between 2021 and 2023 (5,907 to 9,102 diagnoses).

For women, trichomoniasis infection was associated with older age groups (aged 45+, aOR: 2.75 [95% CI:2.53-2.98] vs 15-25yrs). Women self-reporting Black Caribbean ethnicity (aOR: 3.21 [95% CI: 2.95-3.50]) and any other black background (aOR: 3.75 [95% CI:3.25-4.34]) had higher odds of trichomoniasis diagnosis than people of White British ethnicity. Trichomoniasis diagnoses were associated with living in the most deprived areas (vs. least deprived; aOR 3.25 [95% CI: 2.91-3.63]), living in the West Midlands (vs. London; aOR 2.12, [95% CI: 1.96-2.28]) and in Yorkshire and Humber (vs. London; aOR 1.44, [95% CI: 1.31-1.58]). Similar factors were observed to be associated with a diagnosis of trichomoniasis amongst men.

**Discussion:** The targeted screening of women either at high risk of infection (and their partners) or living in areas of higher prevalence, and the introduction of NAAT testing has likely contributed to the increase in diagnoses, especially among Black ethnicities. Although diagnoses have increased in all areas and ethnic groups, the increased likelihood of trichomoniasis among some racially minoritised groups shows the potential effect of compounding sources of health inequity, something which highlights the need for targeted, culturally competent interventions to address this.

## Introduction

Trichomoniasis is a sexually transmitted infection (STI) caused by the flagellated protozoan *Trichomonas vaginalis* (1). Worldwide, trichomoniasis is the most common non-viral STI (2), but prevalence estimates within the UK are low (3). It is estimated that between 50% and 85% of cases are asymptomatic (4–6), symptoms in women include vaginal discharge, vulval itching, dysuria, soreness, swelling and unpleasant odour. Symptoms in men include urethral discharge or dysuria. Men are often diagnosed following contact tracing due to female sexual partners with trichomoniasis (7). If untreated, trichomoniasis may cause perinatal complications (preterm delivery and/or low birthweight infants), infertility (8), epididymitis, decreased sperm motility and prostatitis (9).

The testing policy in the UK’s national trichomoniasis management guidelines (8) is informed by the observed demographic distribution of diagnoses of this STI at sexual health services (SHSs). Historically, in the UK, testing was recommended only in symptomatic women (vulvitis or vaginal discharge) and their sexual partners from the previous 4 weeks (10). More recently, the guidelines were updated to extend recommended screening to asymptomatic women in geographical areas with high diagnosis rates and in women at increased risk (8). An association between trichomoniasis diagnosis and ethnic group has been previously described (3, 11), which informed the recommendation in the 2021 guidelines for asymptomatic testing of Black Caribbean women.

In this analysis of national STI surveillance data, we describe the trends in trichomoniasis diagnoses between 2012 and 2023, and the correlates of diagnoses of this STI in attendees of SHSs in England in 2023.

## Methods

### Study design

The data used for this analysis is from the GUMCAD STI Surveillance System (from now on referred to as ‘GUMCAD’), which includes routinely collected data on attendances at SHSs in England. GUMCAD is an electronic, pseudonymised, individual-level dataset which includes demographic and behavioural data on service attendees and records of all STI diagnoses, including trichomoniasis. Although GUMCAD records tests for most common STIs such as chlamydia and gonorrhoea, it does not include testing data for trichomoniasis over the period considered in this analysis (testing for trichomoniasis was reportable within GUMCAD 2024 onwards).

To cover the two most recent updates of the UK national trichomoniasis management guidelines (in 2014 and 2021-(8, 10)) and associated changes in testing practice, we used data from 1 January 2012 to 31 December 2023 (inclusive) to describe trichomoniasis trends by gender in England. To avoid double-counting only one trichomoniasis diagnosis is counted within a 42-day period as that is the length of an episode of care used for STIs in GUMCAD. This analysis used individual episodes so some individuals can have multiple diagnoses (4% of diagnoses).

### Statistical analysis

We performed a descriptive analysis of socio-demographic and clinical characteristics including age group, ethnicity, world region of birth, Index of Multiple Deprivation (IMD), area of residence, partner notification, HIV status and concurrent infections. Concurrent infection was defined as having any infection within 42 days before or after a trichomoniasis diagnosis with chlamydia, gonorrhoea, first-episode warts or first-episode herpes. Other vaginal conditions were also included in women: bacterial vaginosis and candidiasis.

We excluded consultations that did not have an STI-related need as this analysis is focused on consultations related to testing and diagnosing STIs (12). Consultations solely for reproductive health or HIV care were excluded.

We also calculated the gender-stratified distribution of diagnoses by self-reported ethnic group in 2023 and compared these to the population ethnicity distributions in the 2021 ONS Census data (13).

To assess the correlates of trichomoniasis diagnoses, we performed regression analyses using 2023 data, where the outcome variable was presence or absence of trichomoniasis diagnosis. We excluded attendances occurring at a SHS following partner notification for trichomoniasis as a much higher proportion of these individuals will be tested for trichomoniasis than those attending without partner notification (0.02% [459/2,050,474] attendances among women and 0.13% [1993/1,555,424] attendances among men). As there are multiple attendances for some service users within the time frame of this analysis, we used generalised estimated equations (GEE) with exchangeable working correlation to account for non-independence between attendances by the same user. Different correlation structures were used as sensitivity analysis and similar results were obtained (results not shown). We stratified the analyses by gender due to the differences in testing policy (women are more likely to be tested). Unadjusted and adjusted odds ratios were calculated with 95% confidence intervals.

We included in the model age and ethnicity a priori; other covariates were included following a stepwise approach informed by previous models used in other analyses using GUMCAD data and factors previously known to be associated with trichomoniasis (11). The covariates used in the final model were age group, ethnic group, world region of birth, IMD score and public health region of residence.

All analyses were performed using Stata v18.0 (StataCorp LLC, College Station, TX, USA).

## Results

Between 2012 and 2023 there were 88,861 diagnoses of trichomoniasis in England, 90% of which were in women.

Nationally, trichomoniasis diagnoses increased by 37% between 2012 (6,950 diagnoses) and 2019 (9,490 diagnoses). After a drop in 2020 (5,327 diagnoses) during the first year of the COVID-19 pandemic, diagnoses increased by 54% between 2021 (5,907 diagnoses) and 2023 (9,102 diagnoses) (Figure 1). These 9,102 diagnoses of trichomoniases related to 8,655 people, indicating that there were multiple diagnoses or persistent infections in 398 individuals.

**Figure 1.**
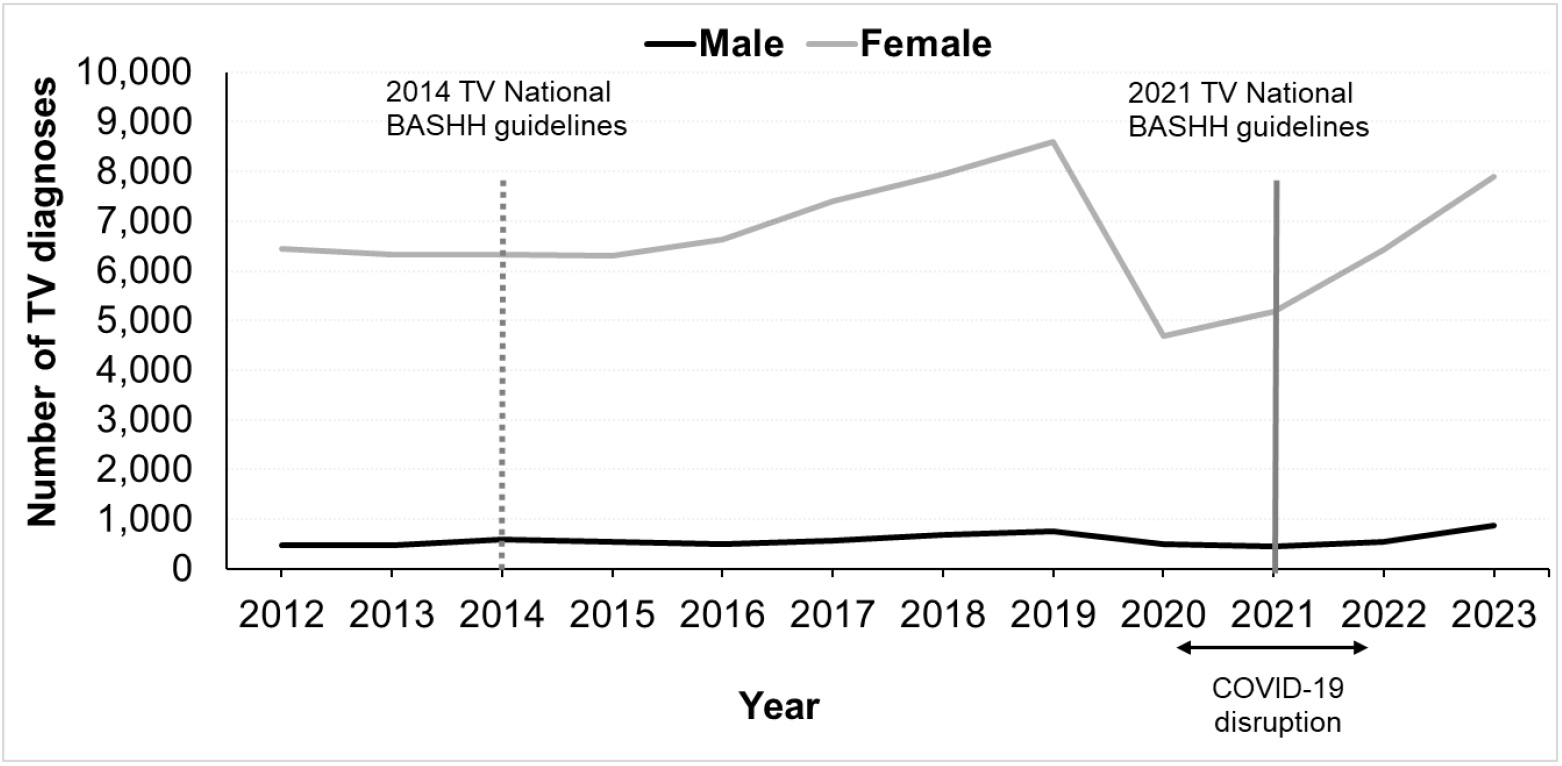
Diagnoses of trichomoniasis, England 2012 to 2023. Footnote: The main changes in the trichomoniasis National BASHH guidelines were as follows: In 2014, introduction of nucleic acid amplification tests and management of infection refractory to first-line treatment. Trichomoniasis testing was recommended in women with symptoms and in men who were contacts of people diagnosed with trichomoniasis and should be considered in those with persistent urethritis. The 2021 guidelines added screening of asymptomatic women in geographical areas of high prevalence and/or in women with associated risk factors (e.g. being a woman of Black Caribbean ethnicity).

The number of trichomoniasis diagnoses was highest in local authority areas in London and the West Midlands (Figure 2). The rates (per 100,000 population) were also highest in these regions (data not shown)

**Figure 2.**
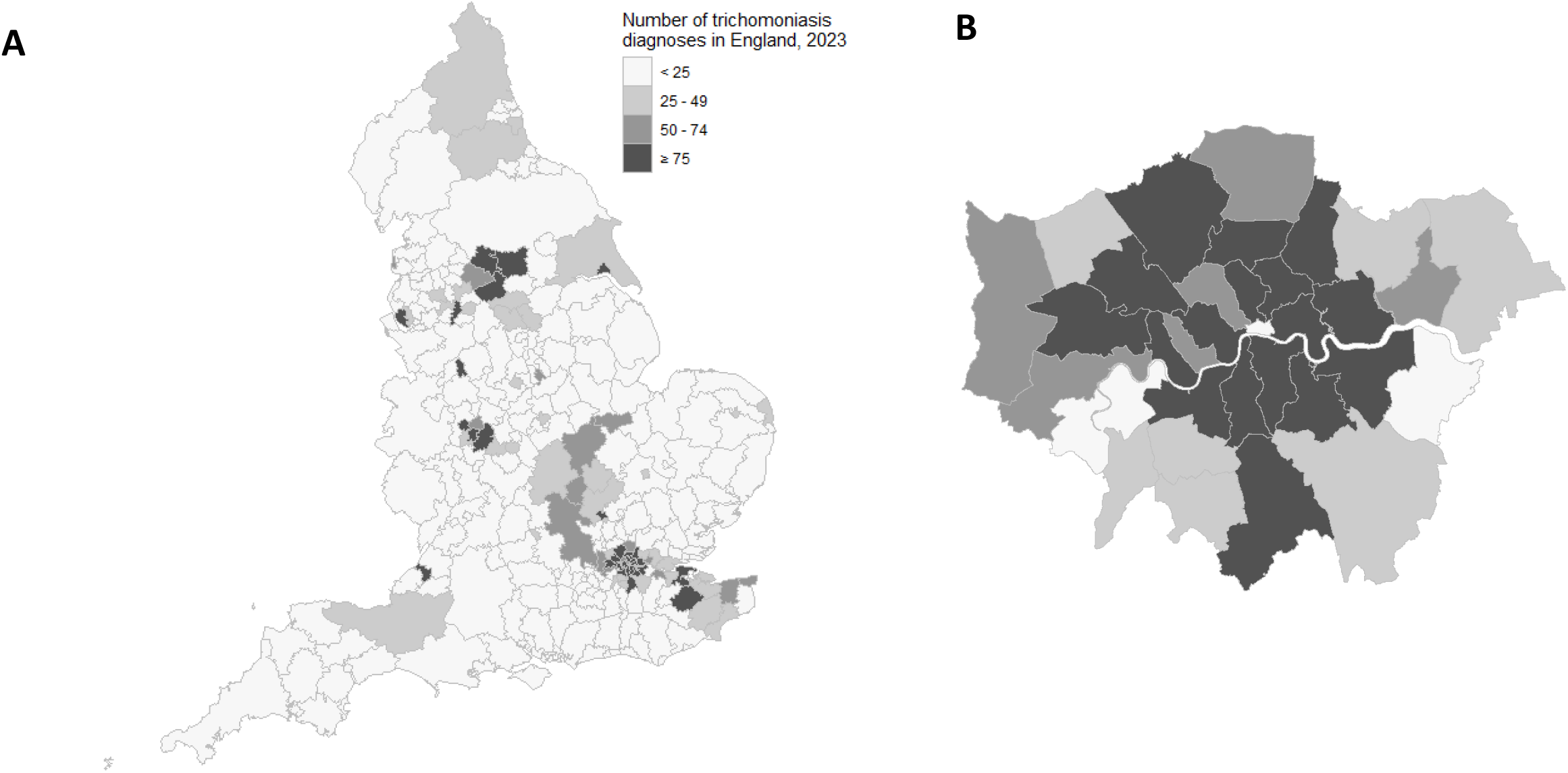
Number of trichomoniasis diagnoses by local authority in England (A) and London (B) 2023.

The number of diagnoses appeared to be increasing among most ethnic groups prior to the COVID-19 pandemic, when there was a decrease in every ethnic group. The number of diagnoses has not recovered to pre-pandemic levels in the majority of groups (Appendix A). Of the total trichomoniasis diagnoses in 2023, 3,937 (43.3%) were in people of White British ethnicity (individuals of this ethnic group make up 73.5% of the population of England) followed by 1,233 diagnoses among people of Black Caribbean ethnicity (13.6% of all diagnoses, despite only being 1.1% of the general population) (Figure 3).

**Figure 3.**
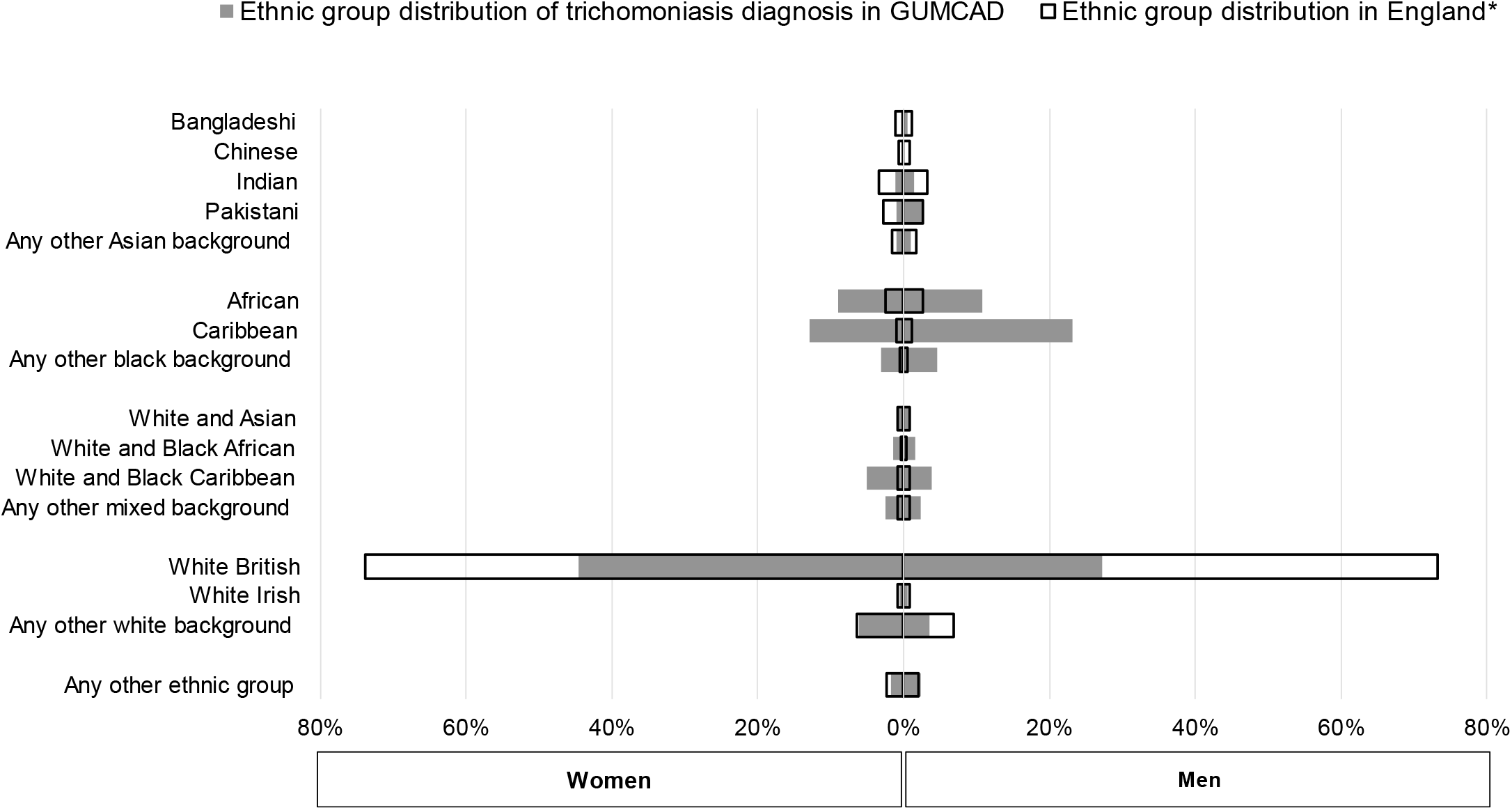
Comparison of the ethnicity distribution among people with trichomoniasis diagnoses and the England population* by gender, England, 2023. *Population source: 2021 ONS Census data by ethnic group. There are inherent limitations when comparing trichomoniasis diagnosis to population data as there is not universal testing for trichomoniasis in England and the asymptomatic testing is restricted to certain demographic groups and geographical locations where they are more likely to reside.

**Figure 4.**
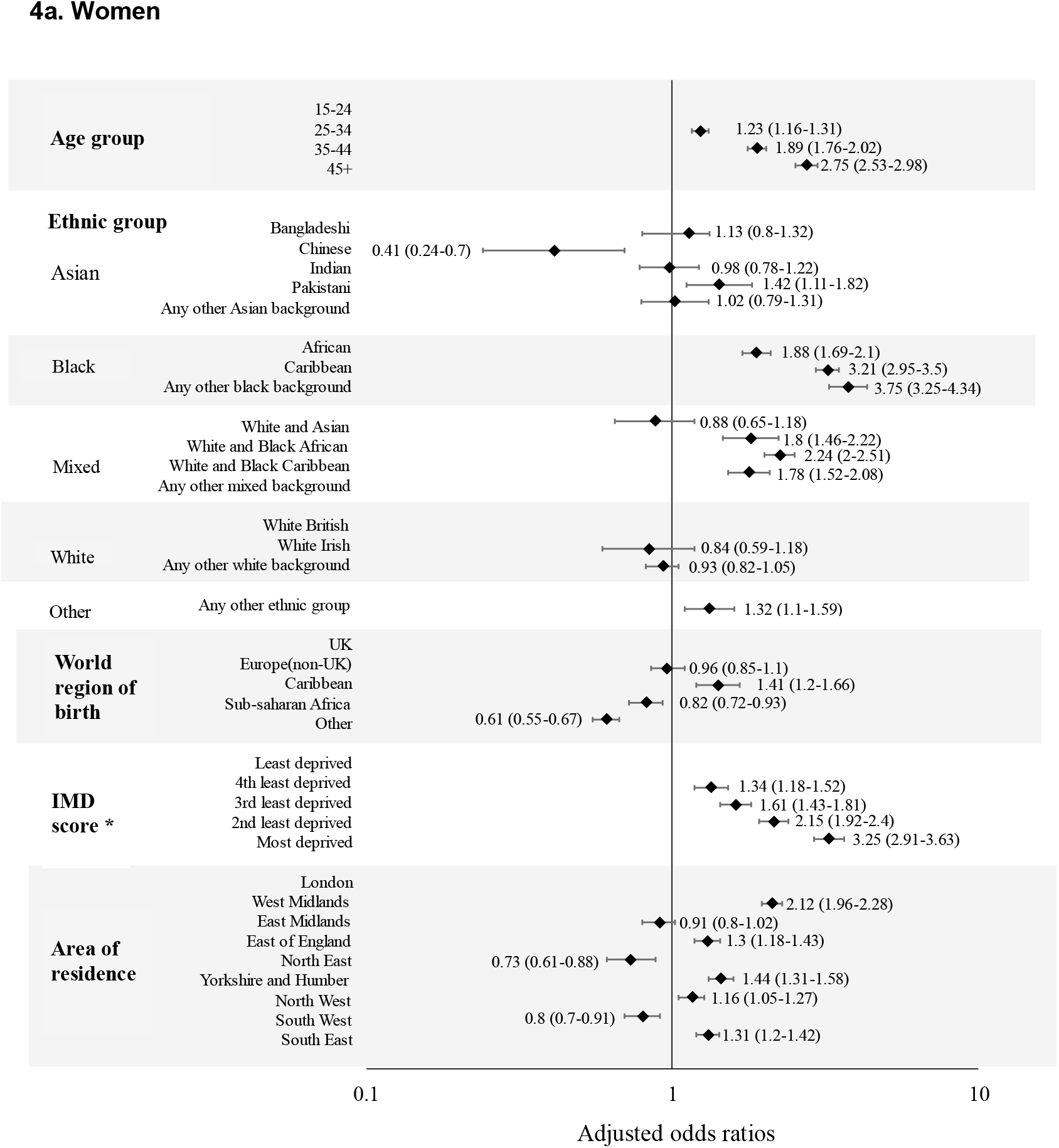
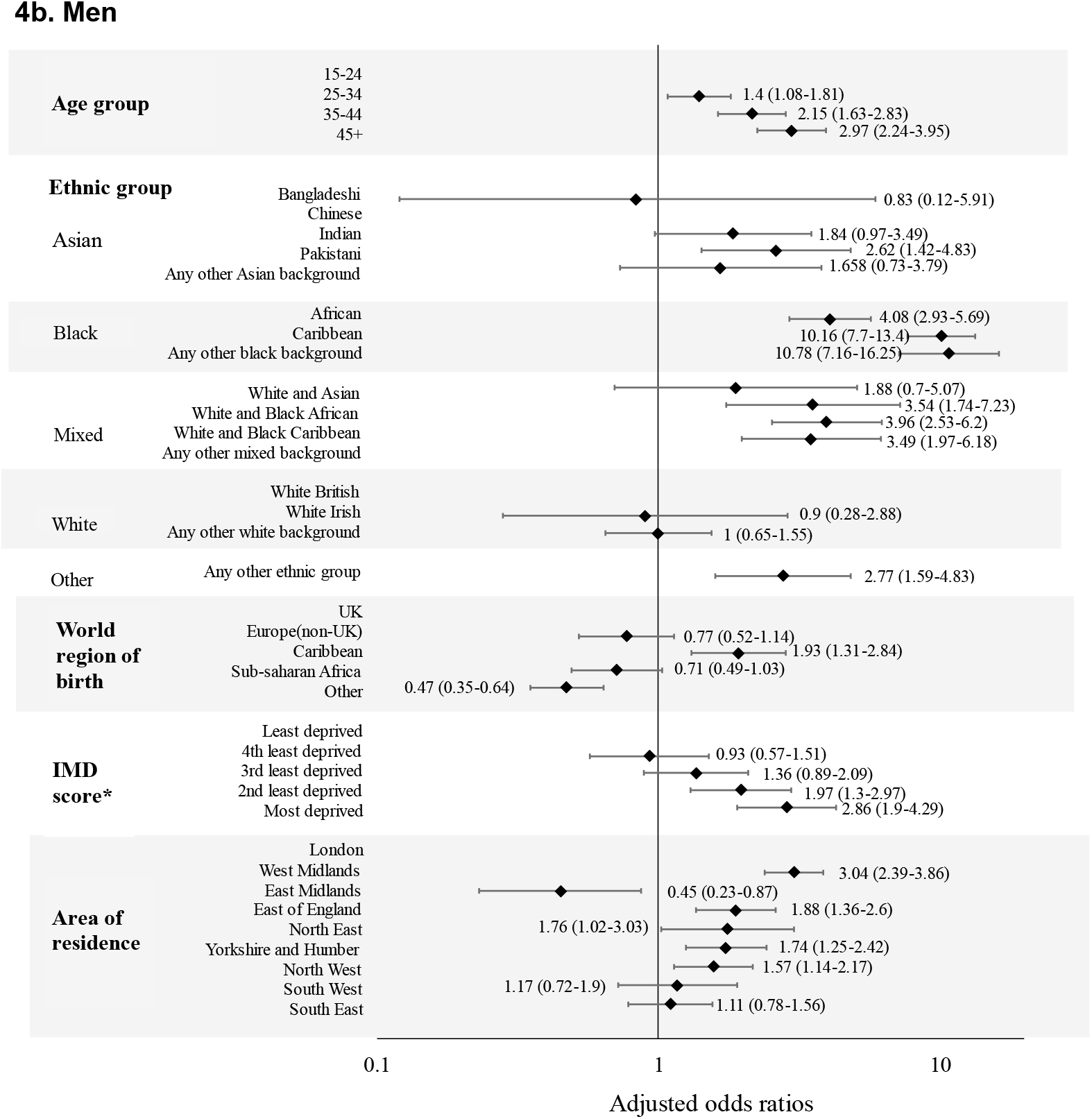
Adjusted associations with trichomoniasis diagnosis among people attending sexual health services in England, by gender, 2023. *IMD: Index of multiple deprivation

### Factors associated with trichomoniasis diagnosis

In univariable analyses (Appendix B), trichomoniasis diagnosis amongst women was associated with age (p<0.001): older women were more likely to have a trichomoniasis diagnosis (uOR for 45+ years vs. 15-25 years: 2.76 [95%CI: 2.56-2.99]). Different ethnic groups experienced different odds of trichomoniasis diagnosis (p<0.001). Women of Black Caribbean ethnicity were 4.11 times (95%CI: 3.81-4.42) and women of Black African ethnicity were 2.06 times (95%CI: 1.89-2.25) more likely to have a trichomoniasis diagnosis than people of White British ethnicity, and those with ‘O’ (A C) were 4.16 times (95%CI: 3.63-4.78) more likely. Those reporting Pakistani ethnicity also had higher odds of diagnosis (uOR: 1.69 [95%CI: 1.32-2.15]). There were also higher odds of trichomoniasis diagnosis in those born in the Caribbean when compared to UK-born individuals (uOR: 4.49 [95% CI: 3.86-5.22]). Trichomoniasis diagnoses were associated with living in the most deprived areas (vs. least deprived; uOR: 3.96 [95%CI: 3.56 – 4.42]) and living in the West Midlands (vs. London; uOR: 1.96 [95%CI: 1.83-2.11]).

Similar factors were observed to be associated with a diagnosis of trichomoniasis amongst men (Appendix C).

Among women, there was an association between trichomoniasis diagnosis and living with HIV (uOR: 1.55 [95% CI: 1.23-1.94]) and having concurrent infections, specifically syphilis (uOR: 5.04 [95%CI: 3.09 – 8.24]) chlamydia (uOR: 3.85 [95%CI: 3.50-4.24]), gonorrhoea (uOR: 3.26 [95%CI: 2.88-3.69]) and herpes (uOR: 1.91 [95%CI: 1.58 - 2.30]). There was also an association with other vaginal conditions such as bacterial vaginosis (uOR: 5.08 [95%CI: 4.72 – 5.47]) and candidiasis (uOR: 2.85 [95%CI: 2.58 – 3.16]).

Among men, there was only an association between trichomoniasis and having concurrent infection with chlamydia (uOR: 2.42 [95%CI: 1.86-3.16]).

In an adjusted model, OR estimates of trichomoniasis diagnosis were similar to the unadjusted estimates. However, there was a decrease in the aOR for women who were Black Caribbean (aOR 3.21 [95%CI: 2.95-3.50]), any other Black background (aOR: 3.75 [95%CI: 3.25-4.34) and for women born in the Caribbean (aOR: 1.41 [95%CI 1.20-1.66]). Following adjustment by sociodemographic factors, we also found that when we compared London region with others in England there was an association (p<0.001) between trichomoniasis diagnosis and living in the West Midlands (aOR: 2.12 [95%CI: 1.96-2.28]), East of England (aOR: 1.30 [95%CI: 1.18-1.43]), North West (aOR: 1.16 [95%CI: 1.05-1.27]), South East (aOR: 1.31 [95%CI: 1.20-1.42]) and Yorkshire and Humber (aOR: 1.44 [95%CI: 1.31-1.58]) among women only.

## Discussion

Using national surveillance data from SHSs in England, we observed an increasing trend in trichomoniasis diagnoses. By 2023 the number of trichomoniasis diagnoses had still not reached pre-COVID-19 levels. However, the number of trichomoniasis diagnoses in 2024 reported by UKHSA in 2025 (10,036 diagnoses) (14) were higher than before the COVID-19 pandemic. In line with the testing policy, most diagnoses were in women and women of Black Caribbean ethnicity had disproportionately high numbers of diagnoses. We also found increased odds of trichomoniasis diagnoses amongst those living in more deprived areas, a pattern which mirrors those previously observed in the epidemiology of both trichomoniasis and other STIs in England (15). People of Black Caribbean ethnicity (and other black ethnicities) are more likely to reside in the most deprived areas compared to other ethnic groups (16–18), so the higher odds of trichomoniasis diagnosis within this population potentially reflect this socioeconomic disparity, as well as the clinical testing policy to test women of this ethnic group residing in those regions (8).

We found a high degree of geographic heterogeneity with more diagnoses in London, the West Midlands, and the North West. This is consistent with the geographic variation and distribution of Black Caribbean ethnicities in England and likely to reflect higher testing in keeping with the current national testing recommendations that were updated in 2021 (8), which advised testing asymptomatic women in areas with higher numbers of diagnoses (for example London and the West Midlands). The increased odds of trichomonas diagnosis in both London and the West Midlands has been recorded previously (11). Additionally, we reported increased odds of diagnosis in the East of England and Yorkshire and Humber for both men and women and in the North West amongst men (although the number of diagnoses remains much lower than women across all regions due to testing guidelines).

Around 4% of those diagnosed with trichomoniasis in 2023 had multiple episodes of trichomoniasis recorded, which could indicate treatment failure or persistent infection. This could potentially be due to metronidazole resistance (19, 20), however data from the UK is currently limited.

These multiple episodes could also be due to reinfection. Partner notification is an important part of STI control and is recommended in the guidelines for Trichomoniasis management (8). In 2023, most diagnoses made following partner notification were among men, which is likely a result of the testing guidelines focusing on women. Only 15% of notified partners were diagnosed with trichomoniasis, which is lower than more commonly reported STIs such as chlamydia and gonorrhoea in England (15). Potential causes of lower proportion of trichomoniasis diagnoses in notified partners could be factors such as the high levels of asymptomatic cases or a lack of awareness of trichomoniasis in the general population reducing drive to receive testing.

The natural history of trichomoniasis in men is poorly understood; recent evidence has shown that spontaneous resolution of infection is less common than previously hypothesised, with 70% of men having a repeat positive test if they had not received treatment (21). It is possible that asymptomatic men may reinfect their previously treated partners. Consequently, partner notification plays an important role in preventing onwards transmission of trichomoniasis and requires services to have the ability for onwards referral when testing needs cannot be met within that service (8, 12).

We found a high level of concurrent infections with other STIs amongst women, particularly bacterial infections such as chlamydia, gonorrhoea, and syphilis. This has been reported in previous epidemiological studies (11). There may be commonality between the sociodemographic risk factors involved in the transmission of both trichomoniasis and other STIs. The groups at increased risk of trichomoniasis previously described also have a high burden of bacterial STI infection, such as Black Caribbean populations and those living in deprived areas(15). One difference found between the epidemiological profile of trichomoniasis and other STIs, is the higher odds of trichomoniasis diagnosis amongst older age groups which differs from chlamydia and gonorrhoea trends in England (15). We also found a bivariate association between trichomoniasis diagnosis and living with HIV. It is possible that increased provision of trichomoniasis testing in HIV care services with a focus on minoritised groups such as Black African and Caribbean individuals would facilitate a more rapid identification of trichomoniasis in these groups with an increased burden (or potentially trichomoniasis diagnosis could lead to more rapid identification of HIV).

### Strengths and limitations

While GUMCAD is a comprehensive dataset of all trichomoniasis diagnoses at SHSs in England, which allows us to present a unique insight into the epidemiology of trichomoniasis, there are multiple limitations to this analysis. One is the limited ability to evaluate the impact of sexual behaviour and whether diagnoses were symptomatic or not, as this information was not available for the period presented in this analysis and is a relatively recent addition to the GUMCAD dataset.

We are also unable to assess test-positivity or testing rates as a trichomoniasis test code was not available in GUMCAD until 2024. This means we cannot distinguish between consultations that had no trichomoniasis diagnosis due to a negative test result versus consultations without a trichomoniasis test, some of whom may be carrying the infection asymptomatically. While it is likely that the increasing trend of trichomoniasis diagnosis is in part due to changes in testing guidelines and latterly a rebound in service provision following the COVID pandemic, it is more difficult to interpret whether an increase in the number of diagnoses may indicate that prevalence of trichomoniasis in England is higher than previously reported. A clinical code to record trichomoniasis testing has been recently introduced in GUMCAD which, in future, can be used to assess test positivity as well as the number of partner contacts who receive screening for trichomoniasis, providing more insight into the burden of this infection in England.

## Conclusion

The epidemiology of trichomoniasis within England has remained similar to previous reports, with higher diagnoses rates amongst women, Black minority ethnic groups and those living in deprived areas. This, and the increasing trend in diagnoses, is likely due to implementation of the updated (2021) trichomoniasis testing guidelines within the UK and the increased accuracy of NAAT testing methods. It is important to continue to be aware of groups at increased risk of trichomoniasis and local areas with increasing diagnoses as this can inform tailored guidance and practice within SHSs and by community partners to improve sexual health. The increased likelihood of trichomoniasis among some racially minoritised groups shows the potential effect of compounding sources of health inequity, something which highlights the need for targeted, culturally competent interventions to address this.

## Acknowledgement

We would like to offer our thanks to all the sexual health services that report GUMCAD data to UKHSA.

## Data availability statement

In its role providing infectious disease surveillance, UK Health Security Agency has permission to handle data obtained through the GUMCAD STI Surveillance System under Regulation 3 of the Health Service (Control of Patient Information) Regulations 2002.

Requests for aggregate data can be made by contacting the GUMCAD, where all publicly released data must adhere to UKHSA data sharing guidelines.

## Author contributions

AKH, GB. HM, SJM and LV curated an analysis plan with support from MC and RPK. AKH and GB carried out the analysis and write-up of the first manuscript draft. All authors contributed to successive drafts and reviewed and approved the final manuscript.

## Ethical approval

All data were collected within statutory approvals granted to the UK Health Security Agency for infectious disease surveillance and control. The UK Health Security Agency has approval to handle data obtained by the GUMCAD STI surveillance system under Regulation 3 of the Health Service (Control of Patient Information) Regulations 2002. Information was held securely and in accordance with the Data Protection Act 2018 and Caldicott guidelines.

## APPENDIX A

**Number of trichomoniasis diagnoses by ethnic group and year, England 2012 to 2023**

**Table A1.**
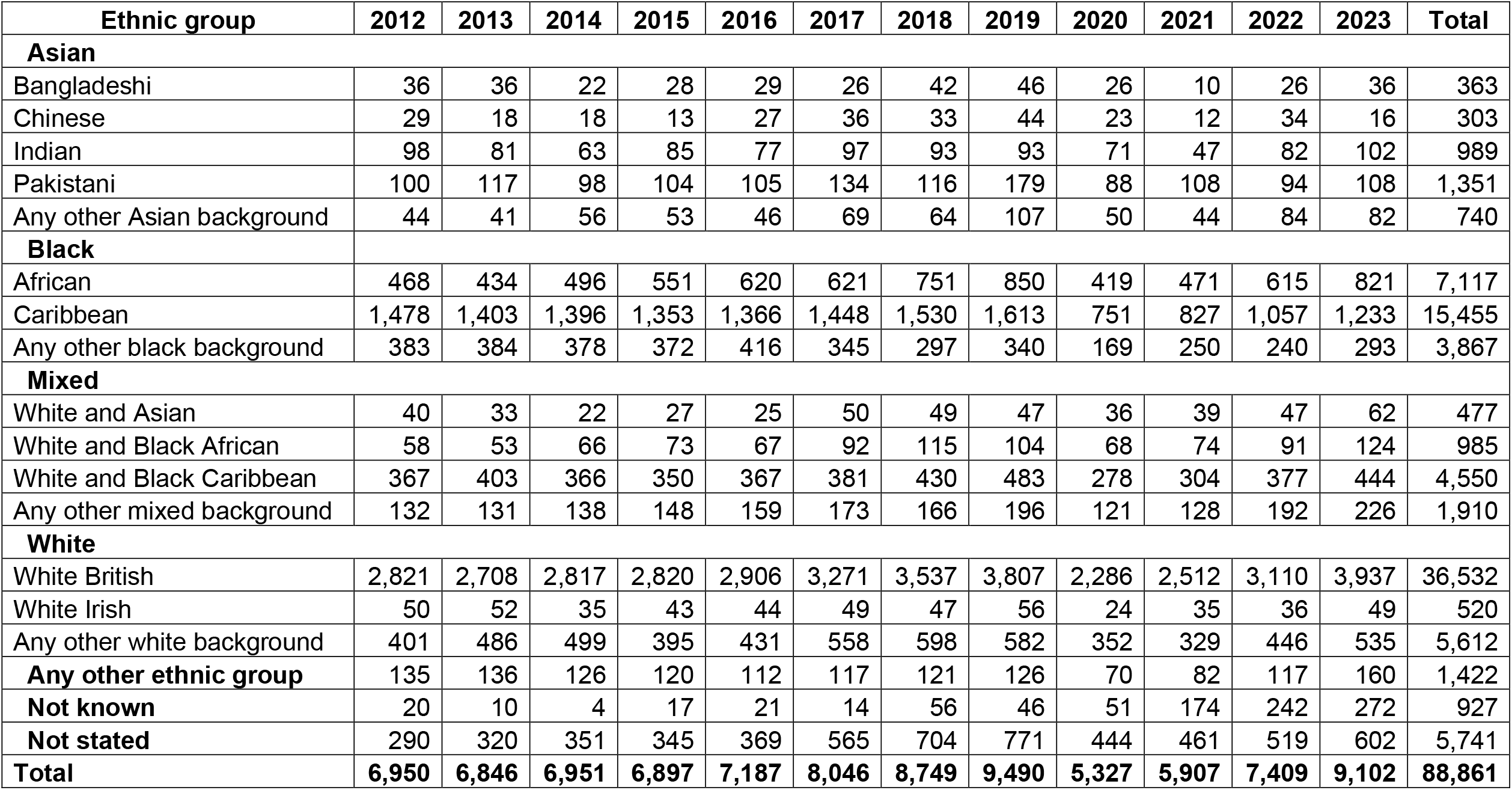

| **Ethnic group** | **2012** | **2013** | **2014** | **2015** | **2016** | **2017** | **2018** | **2019** | **2020** | **2021** | **2022** | **2023** | **Total** |
| --- | --- | --- | --- | --- | --- | --- | --- | --- | --- | --- | --- | --- | --- |
| **Asian** |  |  |  |  |  |  |  |  |  |  |  |  |  |
| Bangladeshi | 36 | 36 | 22 | 28 | 29 | 26 | 42 | 46 | 26 | 10 | 26 | 36 | 363 |
| Chinese | 29 | 18 | 18 | 13 | 27 | 36 | 33 | 44 | 23 | 12 | 34 | 16 | 303 |
| Indian | 98 | 81 | 63 | 85 | 77 | 97 | 93 | 93 | 71 | 47 | 82 | 102 | 989 |
| Pakistani | 100 | 117 | 98 | 104 | 105 | 134 | 116 | 179 | 88 | 108 | 94 | 108 | 1,351 |
| Any other Asian background | 44 | 41 | 56 | 53 | 46 | 69 | 64 | 107 | 50 | 44 | 84 | 82 | 740 |
| **Black** |  |  |  |  |  |  |  |  |  |  |  |  |  |
| African | 468 | 434 | 496 | 551 | 620 | 621 | 751 | 850 | 419 | 471 | 615 | 821 | 7,117 |
| Caribbean | 1,478 | 1,403 | 1,396 | 1,353 | 1,366 | 1,448 | 1,530 | 1,613 | 751 | 827 | 1,057 | 1,233 | 15,455 |
| Any other black background | 383 | 384 | 378 | 372 | 416 | 345 | 297 | 340 | 169 | 250 | 240 | 293 | 3,867 |
| **Mixed** |  |  |  |  |  |  |  |  |  |  |  |  |  |
| White and Asian | 40 | 33 | 22 | 27 | 25 | 50 | 49 | 47 | 36 | 39 | 47 | 62 | 477 |
| White and Black African | 58 | 53 | 66 | 73 | 67 | 92 | 115 | 104 | 68 | 74 | 91 | 124 | 985 |
| White and Black Caribbean | 367 | 403 | 366 | 350 | 367 | 381 | 430 | 483 | 278 | 304 | 377 | 444 | 4,550 |
| Any other mixed background | 132 | 131 | 138 | 148 | 159 | 173 | 166 | 196 | 121 | 128 | 192 | 226 | 1,910 |
| **White** |  |  |  |  |  |  |  |  |  |  |  |  |  |
| White British | 2,821 | 2,708 | 2,817 | 2,820 | 2,906 | 3,271 | 3,537 | 3,807 | 2,286 | 2,512 | 3,110 | 3,937 | 36,532 |
| White Irish | 50 | 52 | 35 | 43 | 44 | 49 | 47 | 56 | 24 | 35 | 36 | 49 | 520 |
| Any other white background | 401 | 486 | 499 | 395 | 431 | 558 | 598 | 582 | 352 | 329 | 446 | 535 | 5,612 |
| **Any other ethnic group** | 135 | 136 | 126 | 120 | 112 | 117 | 121 | 126 | 70 | 82 | 117 | 160 | 1,422 |
| **Not known** | 20 | 10 | 4 | 17 | 21 | 14 | 56 | 46 | 51 | 174 | 242 | 272 | 927 |
| **Not stated** | 290 | 320 | 351 | 345 | 369 | 565 | 704 | 771 | 444 | 461 | 519 | 602 | 5,741 |
| **Total** | **6,950** | **6,846** | **6,951** | **6,897** | **7,187** | **8,046** | **8,749** | **9,490** | **5,327** | **5,907** | **7,409** | **9,102** | **88,861** |

## APPENDIX B

**Risk factors associated with trichomoniasis diagnoses in women attending sexual health services in England, 2023**

**Table B1.**

| Women | Consultations without trichomoniasis diagnosis (%) | Number Trichomoniasis diagnosis (%) | Unadjusted OR (95% CI) | p-value, unadjusted model | Adjusted OR (95% CI) | p-value, adjusted model |
| --- | --- | --- | --- | --- | --- | --- |
| N | 2,031,778 (99.6%) | 7,781 (0.4%) |  |  |  |  |
| **Age group** |  |  |  | <0.001 |  | <0.001 |
| 15-24 | 763,909 (37.6%) | 2,124 (27.3%) | Ref |  | Ref |  |
| 25-34 | 820,206 (40.4%) | 2,865 (36.8%) | 1.27 (1.20-1.35) |  | 1.23 (1.16-1.31) |  |
| 35-44 | 309,145 (15.2%) | 1,726 (22.2%) | 2.02 (1.89-2.15) |  | 1.89 (1.76-2.02) |  |
| 45+ | 138,464 (6.8%) | 1,066 (13.7%) | 2.76 (2.56-2.99) |  | 2.75 (2.53-2.98) |  |
| **Ethnic group** |  |  |  |  |  |  |
| **Asian** |  |  |  | <0.001 |  | <0.001 |
| Bangladeshi | 8,716 (0.4%) | 30 (0.4%) | 1.2 (0.82-1.76) |  | 1.13 (0.8-1.32) |  |
| Chinese | 18,476 (0.9%) | 16 (0.2%) | 0.3 (0.18-0.49) |  | 0.41 (0.24-0.7) |  |
| Indian | 32,700 (1.6%) | 85 (1.1%) | 0.89 (0.71-1.11) |  | 0.98 (0.78-1.22) |  |
| Pakistani | 16,531 (0.8%) | 82 (1.1%) | 1.69 (1.32-2.15) |  | 1.42 (1.11-1.82) |  |
| Any other Asian background | 31,564 (1.6%) | 73 (0.9%) | 0.79 (0.62-1.00) |  | 1.02 (0.81-1.31) |  |
| **Black** |  |  |  |  |  |  |
| African | 117,546 (5.8%) | 697 (9.0%) | 2.06 (1.89-2.25) |  | 1.88 (1.69-2.1) |  |
| Caribbean | 87,924 (4.3%) | 996 (12.8%) | 4.11 (3.81-4.42) |  | 3.21 (2.95-3.5) |  |
| Any other black background | 20,410 (1.0%) | 248 (3.2%) | 4.16 (3.63-4.78) |  | 3.75 (3.25-4.34) |  |
| **Mixed** |  |  |  |  |  |  |
| White and Asian | 22,928 (1.1%) | 51 (0.7%) | 0.77 (0.58-1.03) |  | 0.88 (0.65-1.18) |  |
| White and Black African | 20,761 (1.0%) | 106 (1.4%) | 1.78 (1.45-2.18) |  | 1.8 (1.46-2.22) |  |
| White and Black Caribbean | 54,561 (2.7%) | 398 (5.1%) | 2.56 (2.29-2.85) |  | 2.24 (2-2.51) |  |
| Any other mixed background | 41,887 (2.1%) | 194 (2.5%) | 1.61 (1.38-1.88) |  | 1.78 (1.52-2.08) |  |
| **White** |  |  |  |  |  |  |
| White British | 1,192,587 (58.7%) | 3,461 (44.5%) | Ref |  | Ref |  |
| White Irish | 22,514 (1.1%) | 43 (0.6%) | 0.66 (0.47-0.91) |  | 0.84 (0.59-1.18) |  |
| Any other white background | 186,590 (9.2%) | 474 (6.1%) | 0.87 (0.79-0.96) |  | 0.93 (0.82-1.05) |  |
| **Other** |  |  |  |  |  |  |
| Any other ethnic group | 39,380 (1.9%) | 135 (1.7%) | 1.2 (1.01-1.43) |  | 1.32 (1.1-1.59) |  |
| **Not known** | 20,436 (1.0%) | 228 (2.9%) | 3.73 (3.25-4.28) |  | 3.57 (3.09-4.12) |  |
| **Not stated** | 96,267<br>(4.7%) | 464 (6.0%) | 1.59 (1.43-1.76) |  | 2.17 (1.91-2.47) |  |
| **World region of birth** |  |  |  | <0.001 |  | <0.001 |
| UK | 1,520,825<br>(74.9%) | 5,804<br>(74.6%) | Ref |  | Ref |  |
| Europe(non-UK) | 129,452<br>(6.4%) | 435 (5.6%) | 0.87 (0.79-0.96) |  | 0.96 (0.85-1.1) |  |
| Caribbean | 11,234<br>(0.6%) | 188 (2.4%) | 4.49 (3.86-5.22) |  | 1.41 (1.20-1.66) |  |
| Sub-saharan Africa | 67,103<br>(3.3%) | 433 (5.6%) | 1.66 (1.5-1.84) |  | 0.82 (0.72-0.93) |  |
| Other | 303,164<br>(14.9%) | 921 (11.8%) | 0.77 (0.72-0.83) |  | 0.61 (0.55-0.67) |  |
| **IMD score\*\*** |  |  |  | <0.001 |  | <0.001 |
| Least deprived | 233,523<br>(11.8%) | 393 (5.2%) | Ref |  | Ref |  |
| 4th least deprived | 314,103<br>(15.9%) | 707 (9.4%) | 1.34 (1.18-1.53) |  | 1.34 (1.18-1.52) |  |
| 3rd least deprived | 413,246<br>(20.9%) | 1,149<br>(15.3%) | 1.67 (1.48-1.88) |  | 1.61 (1.43-1.81) |  |
| 2nd least deprived | 539,838<br>(27.3%) | 2,110<br>(28.1%) | 2.36 (2.11-2.64) |  | 2.15 (1.92-2.4) |  |
| Most deprived | 476,862<br>(24.1%) | 3,146<br>(41.9%) | 3.96 (3.56-4.42) |  | 3.25 (2.91-3.63) |  |
| **Area of residence** |  |  |  | <0.001 |  | <0.001 |
| London | 758,132<br>(37.3%) | 2,849<br>(36.6%) | Ref |  | Ref |  |
| West Midlands | 150,326<br>(7.4%) | 1,144<br>(14.7%) | 1.96 (1.83-2.11) |  | 2.12 (1.96-2.28) |  |
| East Midlands | 121,527<br>(6.0%) | 331 (4.3%) | 0.7 (0.62-0.78) |  | 0.91 (0.80-1.02) |  |
| East of England | 165,808<br>(8.2%) | 594 (7.6%) | 0.92 (0.84-1.01) |  | 1.30 (1.18-1.43) |  |
| North East | 60,503<br>(3.0%) | 130 (1.7%) | 0.56 (0.47-0.67) |  | 0.73 (0.61-0.88) |  |
| Yorkshire and Humber | 154,984<br>(7.6%) | 675 (8.7%) | 1.13 (1.03-1.23) |  | 1.44 (1.31-1.58) |  |
| North West | 180,328<br>(8.9%) | 658 (8.5%) | 0.95 (0.87-1.03) |  | 1.16 (1.05-1.27) |  |
| South West | 143,602<br>(7.1%) | 280 (3.6%) | 0.5 (0.44-0.56) |  | 0.80 (0.70-0.91) |  |
| South East | 254,104<br>(12.5%) | 907 (11.7%) | 0.89 (0.83-0.96) |  | 1.31 (1.20-1.42) |  |
| Not Known | 42,464<br>(2.1%) | 213 (2.7%) | 1.29 (1.12-1.49) |  | 0 (0-0) |  |
| **HIV status** |  |  |  | <0.001 |  |  |
| Negative/unknown | 2,017,673<br>(99.3%) | 7,694<br>(98.9%) | Ref |  |  |  |
| Person living with HIV | 14,105<br>(0.7%) | 87 (1.1%) | 1.55 (1.23-1.94) |  |  |  |
| **Concurrent infections** |  |  |  |  |  |  |
| Chlamydia | 33,777<br>(1.7%) | 499 (6.4%) | 3.85 (3.50-4.24) | <0.001 |  |  |
| Gonorrhoea | 21,257<br>(1.0%) | 265 (3.4%) | 3.26 (2.88-3.69) | <0.001 |  |  |
| Syphilis | 957 (0.0%) | 21 (0.3%) | 5.04 (3.09-8.24) | <0.001 |  |  |
| Herpes | 16,188<br>(0.8%) | 125 (1.6%) | 1.91 (1.58-2.30) | <0.001 |  |  |
| Warts | 9,869 (0.5%) | 40 (0.5%) | 1.03 (0.75-1.41) | 0.793 |  |  |
| **Vaginal conditions** |  |  |  |  |  |  |
| Bacterial Vaginosis | 51,624 (2.5%) | 991 (12.7%) | 5.08 (4.72-5.47) | <0.001 |  |  |
| candidiasis | 42,064 (2.1%) | 493 (6.3%) | 2.85 (2.58-3.16) | <0.001 |  |  |

## APPENDIX C

**Risk factors associated with trichomoniasis diagnosis in men attending sexual health services in England, 2023**

**Table C1.**

Appendix C. Risk factors associated with trichomoniasis diagnosis in men attending sexual health services in England, 2023
| Men | Consultations without trichomoniasis diagnoses (%) | Number Trichomoniasis diagnoses (%) | Unadjusted OR (95% CI) | p-value, unadjusted model | Adjusted OR (95% CI) | p-value, adjusted model |
| --- | --- | --- | --- | --- | --- | --- |
| N | 1,545,475 (99.96%) | 602 (0.04%) |  |  |  |  |
| **Age group** |  |  |  | <0.001 |  | <0.001 |
| 15-24 | 353,862 (22.9%) | 89 (14.8%) | Ref |  | Ref |  |
| 25-34 | 624,531 (40.4%) | 205 (34.1%) | 1.32 (1.02-1.70) |  | 1.40 (1.08-1.81) |  |
| 35-44 | 319,608 (20.7%) | 156 (25.9%) | 1.96 (1.50-2.56) |  | 2.15 (1.63-2.83) |  |
| 45+ | 247,429 (16.0%) | 152 (25.2%) | 2.46 (1.88-3.23) |  | 2.97 (2.24-3.95) |  |
| **Ethnic group** |  |  |  |  |  |  |
| **Asian** |  |  |  | <0.001 |  | <0.001 |
| Bangladeshi | 7,255 (0.5%) | 1 (0.2%) | 0.70 (0.1-4.99) |  | 0.83 (0.12-5.91) |  |
| Chinese | 20,099 (1.3%) | 0 (0.0%) |  |  | 1.00 (0.00-0.00) |  |
| Indian | 35,147 (2.3%) | 10 (1.7%) | 1.43 (0.76-2.72) |  | 1.84 (0.97-3.49) |  |
| Pakistani | 19,902 (1.3%) | 13 (2.2%) | 3.31 (1.88-5.84) |  | 2.62 (1.42-4.83) |  |
| Any other Asian background | 32,458 (2.1%) | 7 (1.2%) | 1.12 (0.52-2.39) |  | 1.66 (0.73-3.79) |  |
| **Black** |  |  |  |  |  |  |
| African | 89,625 (5.8%) | 66 (11.0%) | 3.76 (2.82-5.03) |  | 4.08 (2.93-5.69) |  |
| Caribbean | 57,430 (3.7%) | 140 (23.3%) | 12.50 (9.90-15.78) |  | 10.16 (7.70-13.40) |  |
| Any other black background | 15,682 (1.0%) | 29 (4.8%) | 9.44 (6.35-14.03) |  | 10.78 (7.16-16.25) |  |
| **Mixed** |  |  |  |  |  |  |
| White and Asian | 15,123 (1.0%) | 4 (0.7%) | 1.35 (0.50-3.64) |  | 1.88 (0.70-5.07) |  |
| White and Black African | 14,449 (0.9%) | 8 (1.3%) | 2.84 (1.39-5.77) |  | 3.54 (1.74-7.23) |  |
| White and Black Caribbean | 30,218 (2.0%) | 23 (3.8%) | 3.89 (2.51-6.03) |  | 3.96 (2.53-6.20) |  |
| Any other mixed background | 31,547 (2.0%) | 15 (2.5%) | 2.46 (1.40-4.30) |  | 3.49 (1.97-6.18) |  |
| **White** |  |  |  |  |  |  |
| White British | 830,885 (53.8%) | 163 (27.1%) | Ref |  | Ref |  |
| White Irish | 24,392 (1.6%) | 4 (0.7%) | 0.82 (0.24-2.77) |  | 0.90 (0.28-2.88) |  |
| Any other white background | 190,892 (12.4%) | 26 (4.3%) | 0.70 (0.46-1.06) |  | 1.00 (0.65-1.55) |  |
| **Other** |  |  |  |  |  |  |
| Any other ethnic group | 40,327 (2.6%) | 14 (2.3%) | 1.79 (1.04-3.10) |  | 2.77 (1.59-4.83) |  |
| **Not known** | 15,904 (1.0%) | 32 (5.3%) | 10.11 (6.85-14.94) |  | 10.8 (7.14-16.35) |  |
| **Not stated** | 74,140 (4.8%) | 47 (7.8%) | 3.18 (2.27-4.45) |  | 5.72 (3.85-8.49) |  |
| **World region of birth** |  |  |  | <0.001 |  | <0.001 |
| UK | 1,081,643 (70.0%) | 412 (68.4%) | Ref |  | Ref |  |
| Europe(non-UK) | 132,410 (8.6%) | 29 (4.8%) | 0.98 (0.61-1.58) |  | 0.77 (0.52-1.14) |  |
| Caribbean | 9,558 (0.6%) | 43 (7.1%) | 1.58 (1.03-2.41) |  | 1.93 (1.31-2.84) |  |
| Sub-Saharan Africa | 57,871 (3.7%) | 41 (6.8%) | 2.47 (1.67-3.67) |  | 0.71 (0.49-1.03) |  |
| Other | 263,993 (17.1%) | 77 (12.8%) | 4.67 (3.18-6.86) |  | 0.47 (0.35-0.64) |  |
| **IMD score\*\*** |  |  |  | <0.001 |  | <0.001 |
| Least deprived | 174,467 (11.6%) | 29 (5.0%) | Ref |  | Ref |  |
| 4th least deprived | 240,489 (16.0%) | 39 (6.7%) | 0.98 (0.61-1.58) |  | 0.93 (0.57-1.51) |  |
| 3rd least deprived | 322,129 (21.5%) | 84 (14.4%) | 1.58 (1.03-2.41) |  | 1.36 (0.89-2.09) |  |
| 2nd least deprived | 432,671 (28.8%) | 176 (30.2%) | 2.47 (1.67-3.67) |  | 1.97 (1.30-2.97) |  |
| Most deprived | 330,118 (22.0%) | 254 (43.6%) | 4.67 (3.18-6.86) |  | 2.86 (1.90-4.29) |  |
| **Area of residence** |  |  |  | <0.001 |  | <0.001 |
| London | 672,555 (43.5%) | 249 (41.4%) | Ref |  | Ref |  |
| West Midlands | 98,654 (6.4%) | 107 (17.8%) | 2.87 (2.28-3.62) |  | 3.04 (2.39-3.86) |  |
| East Midlands | 76,439 (4.9%) | 10 (1.7%) | 0.34 (0.18-0.64) |  | 0.45 (0.23-0.87) |  |
| East of England | 122,984 (8.0%) | 52 (8.6%) | 1.12 (0.82-1.53) |  | 1.88 (1.36-2.60) |  |
| North East | 41,226 (2.7%) | 14 (2.3%) | 0.89 (0.52-1.53) |  | 1.76 (1.02-3.03) |  |
| Yorkshire and Humber | 103,372 (6.7%) | 47 (7.8%) | 1.22 (0.88-1.68) |  | 1.74 (1.25-2.42) |  |
| North West | 119,371 (7.7%) | 46 (7.6%) | 1.02 (0.74-1.40) |  | 1.57 (1.14-2.17) |  |
| South West | 96,319 (6.2%) | 21 (3.5%) | 0.57 (0.36-0.91) |  | 1.17 (0.72-1.90) |  |
| South East | 179,181 (11.6%) | 42 (7.0%) | 0.62 (0.45-0.86) |  | 1.11 (0.78-1.56) |  |
| Not Known | 35,374 (2.3%) | 14 (2.3%) | 1.05 (0.61-1.80) |  | 0 (0-0) |  |
| **HIV status** |  |  |  |  |  |  |
| Negative/unknown | 1,488,352 (96.3%) | 591 (98.2%) | Ref |  |  |  |
| Person living with HIV | 57,123 (3.7%) | 11 (1.8%) | 0.49 (0.27-0.88) | 0.018 |  |  |
| **Concurrent infections** |  |  |  |  |  |  |
| Chlamydia | 45,722 (3.0%) | 48 (8.0%) | 2.42 (1.86-3.16) | <0.001 |  |  |
| Gonorrhoea | 60,233 (3.9%) | 21 (3.5%) | 0.72 (0.48-1.05) | 0.625 |  |  |
| Syphilis | 8,179 (0.5%) | 0 (0.0%) |  |  |  |  |
| Herpes | 9,181 (0.6%) | 2 (0.3%) | 0.38 (0.09-1.51) | 0.390 |  |  |
| Warts | 14,566 (0.9%) | 4 (0.7%) | 0.60 (0.25-1.43) | 0.454 |  |  |

